# Point-of-care electroencephalography for prediction of postoperative delirium in older adults undergoing elective surgery: protocol for a prospective cohort study

**DOI:** 10.1101/2025.10.20.25338386

**Authors:** Vikas N. Vattipally, Patrick Kramer, Nada Abouelseoud, Isha Yeleswarapu, A. Daniel Davidar, Joseph M. Dardick, Ali Bydon, Timothy F. Witham, Daniel Lubelski, Kathryn Rosenblatt, Judy Huang, Chetan Bettegowda, Frederick Sieber, Esther S. Oh, Sridevi V. Sarma, Ozan Akca, Nicholas Theodore, Tej D. Azad

**Author notes:** equal contribution (Nicholas Theodore, Tej D. Azad). **Correspondence:** Tej D. Azad, MD, MS Department of Neurosurgery Johns Hopkins Hospital 1800 Orleans Street Baltimore, MD 21287.

## Abstract

**Background:** Postoperative delirium (POD) is a complication of surgery in older adults associated with adverse outcomes. Current screening methods demonstrate poor interrater reliability, and conventional electroencephalography (EEG)-based screening requires intensive setup. Point-of-care (POC) EEG technology offers a rapid and objective alternative that may capture neurophysiological signatures of delirium risk. When combined with baseline and perioperative variables, POC EEG may enable prediction of POD before clinical manifestation. In this study, we aim to develop a POD prediction model using POC EEG as well as explore secondary outcomes such as longer-term cognitive impairment and postoperative pain.

**Methods:** This is a prospective cohort study enrolling older adults (≥60 years) undergoing elective non-cranial inpatient surgery at two academic hospitals. The target cohort size is 150 participants, determined by an events-per-parameter approach. All participants undergo baseline cognitive testing and pain assessment using the Montreal Cognitive Assessment (MoCA) and Numeric Rating Scale. The primary outcome is POD, while secondary outcomes include follow-up MoCA scores and postoperative pain scores. POD is assessed immediately after surgery and every 12 hours during the admission with the 4AT tool. Perioperative EEG is acquired using the Ceribell EEG system (Ceribell, Inc.) across standardized preoperative, intraoperative, and postoperative phases. EEG features such as spectral power, alpha/delta ratio, and burst suppression ratio are analyzed in relation to outcomes. Predictive models will be developed using regularized logistic regression with nested feature sets and model performance will be evaluated.

**Discussion:** This study evaluates whether POC EEG can accurately predict POD in older adults undergoing elective surgery, as well as longer-term cognitive impairment and postoperative pain. This approach could enable early identification of high-risk patients and facilitate targeted preventive strategies. By generating a validated risk model, multimodal exploratory analyses, and openly available datasets, this work aims to advance the practical management of perioperative outcomes.

**Registration:** None.

**Strengths and Limitations:**

- Prospective, multicenter design with standardized perioperative EEG acquisition.
- Use of an FDA-cleared, rapid-deployment EEG device ensures clinical feasibility.
- Integration of EEG, cognitive, and biomarker data enables multimodal prediction.
- Modest single-institution sample size may limit generalizability.
- Limited-channel EEG montage may reduce spatial resolution and signal detail.

## BACKGROUND

Delirium is defined as an acute alteration of consciousness, cognition, and attention that fluctuates in severity.^1^ Postoperative delirium (POD) is a common and diagnostically challenging complication of surgery in older adults.^2^ Its incidence varies widely, ranging from 5-65% depending on the type of surgery.^3^ Despite its prevalence, delirium remains underrecognized in up to two-thirds of affected patients due to its heterogeneous manifestations, symptom overlap with some neurocognitive disorders, and fluctuating nature.^4,5^ This diagnostic gap has clinical consequences, as POD is associated with increased short-term and long-term mortality risk per delirium day, persistent functional decline, and longer hospitalization.^4–6^ Additionally, POD contributes to an estimated adjusted cost of $44,291 per patient within the first postoperative year and exceeds $4 billion in aggregate healthcare expenditures.^4,8^

The underdiagnosis of POD reflects systemic deficiencies in implementation rather than inherent flaws in available tools. Widely adopted instruments such as the Confusion Assessment Method (CAM) demonstrate robust sensitivity (94%) and specificity (89%) in controlled research environments.^9^ However, real-world clinical administration of the CAM demonstrates wide variability in sensitivity (25-86%) and inconsistent interrater reliability based on the expertise of the assessor.^10^ Furthermore, some conventional POD diagnostic tools lack adaptability for serial monitoring and may be susceptible to practice effects. Diagnostic specificity further declines in patients with preexisting neurocognitive conditions such as dementia due to symptom overlap.^10^ Consequently, low adherence to standardized screening protocols and variability in real-world administration likely contribute to persistent underrecognition and suboptimal management of POD.^11^

The limitations of traditional POD screening methods call for a quantifiable and administrator-independent diagnostic tool. Conventional electroencephalography (EEG) is capable of not only detecting but also predicting the onset of an altered cognitive state.^12–14^ Standard clinical EEG reveals well-characterized patterns in delirium such as changes in alpha power (8-13 Hz) and a lower degree of delta variability (1-4 Hz).^15,16^ However, its use as a screening tool is limited by equipment size, need for trained staff, and the requirement for expert review, which restricts rapid deployment in clinical settings. A possible way to utilize the benefit of EEG technologies while overcoming their limitations is through point-of-care (POC) EEG (*i.e.,* headband-based) with a handheld recorder. Such technologies can overcome the complexity of conventional EEG setup while still providing quantifiable signals of cerebral dysfunction, minimizing comorbidity-related confounds, and avoiding administrator-dependent results. Additionally, when coupled with other potential indicators of delirium such as blood-based biomarkers,^17^ these POC EEG signals could be used to develop a machine learning-powered model that offers a standardized and easy-to-use approach to predict the onset of delirium.

Replacing subjective tools with EEG- and blood biomarker-based diagnostics may enhance diagnostic precision, promote earlier intervention, and reduce both patient-level and system-level consequences of POD. A transformative approach must therefore not only improve detection but also enable early prediction. Identifying patients at high risk for delirium before its full clinical manifestation provides a critical window for preemptive intervention, which may mitigate its severity or prevent its onset altogether. Accordingly, the aim of this prospective cohort study is to develop and evaluate a POC EEG-based machine learning model to predict POD incidence among older adult (≥60 years) patients undergoing elective surgery, incorporating baseline and perioperative variables such as blood biomarkers in nested blocks as adjunctive predictive features. Additionally, we seek to investigate associations of these perioperative features with secondary outcomes such as longer-term cognitive impairment and postoperative pain. We hypothesize that POC EEG monitoring is capable of providing reproducible waveform patterns to predict POD when combined with powerful machine learning analytic techniques. The paradigm shift from delirium detection to prediction that our study would allow, in addition to potential prognostication for secondary outcomes, has the potential to improve outcomes for patients undergoing surgery.

## METHODS AND DESIGN

This is a prospective observational cohort study that has been approved by the Johns Hopkins University Institutional Review Board (IRB) (IRB00463945). All study procedures adhere to the Transparent Reporting of a multivariable prediction model for Individual Prognosis or Diagnosis (TRIPOD+AI).^18^ Patients’ verbal and written informed consents are obtained before their inclusion in the study, and all participants are given the opportunity to exit the study at any time. Data were stored in an IRB-approved REDCap (Nashville, TN) directory and an online, encrypted portal hosted by the sponsoring EEG company. Patient data was also extracted from our institution’s electronic health record (Epic Systems, Verona, WI). This study is classified as non-significant risk (NSR) by our IRB. Neither patients nor the public were not involved with the design of this study.

### Study Setting and Participants

This study is being conducted between two hospitals (Johns Hopkins Hospital and Johns Hopkins Bayview Medical Center) within our health system located in Baltimore, Maryland, United States. We are prospectively enrolling older adult patients (≥60 years) undergoing inpatient elective surgery and began enrollment in August 2025. We selected this age cutoff instead of the more common 65 year (*i.e.* Medicare eligibility age) to optimize the balance between sensitivity and specificity for detecting POD while avoiding unnecessary exclusion of the substantial number of surgical patients aged 60-64 treated at our institution. Patients are excluded at screening if they are undergoing a surgery involving the cranium, have a history of prior cranial surgery or a condition affecting baseline cognitive or neurological function (*i.e.*, Alzheimer’s disease, epilepsy), or take a medication known to alter mental status (*i.e.,* ketamine, opioids) at baseline. These exclusions are being performed to prevent intraoperative physical interference from the EEG device, reduce potential signal artifact related to prior operations, and lower confounding of delirium incidence from baseline cognitive factors. Enrollment is actively underway, and we aim to recruit approximately 150 patients as per the following events-per-parameter (EPP) approach:^19^

*(4 parameters) x (5 EPP) = (20 events) / (0.13 events/participant)* ≈ *150 participants*

Equation 1. Determination of target cohort size.

We identified the following parameters for potential inclusion in our analysis: age, perioperative benzodiazepine exposure and dosing, and 2-3 EEG-derived parameters. Based on the exploratory nature of our model and a limited number of predictors, we conservatively used an EPP of 5 for initial estimation. The incidence of POD was set at 13% for this calculation based on prior data in older adults undergoing elective spinal surgery,^20^ as this population was prioritized for initial enrollment due to established collaborations between our study team and spine surgery faculty at our institution. If the observed incidence of POD is less than 10%, we will extend enrollment to a maximum of 200 patients to ensure an adequate number of events.

### Baseline Measurements

Following informed consent, all participants undergo a standardized baseline evaluation. Cognitive status is assessed using the Montreal Cognitive Assessment (MoCA), which provides a quantitative measure of global cognition and serves as a reference for subsequent postoperative assessments.^21^ Subjective pain scores are also recorded after prompting the patient to rate their current pain on the Numeric Rating Scale (NRS) from 0 (no pain) to 10 (most pain). Next, peripheral blood (10 mL) will be collected into cell-free DNA (cfDNA) collection tubes (Streck, La Vista, NE),^22^ processed by centrifugation, and stored for biomarker analyses within 48 hours of collection.

### EEG Device and Acquisition Schedule

Perioperative EEG is acquired using the Ceribell EEG System, produced by Ceribell, Inc. (Sunnyvale, CA). We have entered a research agreement with the company to loan two recording devices free-of-charge, but the company is not involved in study design or analysis.^23^ This FDA-cleared device consists of a compact handheld EEG recorder and a disposable ten-electrode, eight-channel flexible headband, which is available in two sizes based on the patient’s head circumference **(Figure 1)**. The system is designed for rapid deployment in clinical environments, with a setup time of less than five minutes, and can operate for approximately 24 hours on a single battery charge.^24^ Data are transmitted securely to a cloud-based platform that enables storage, review, and analysis in near real-time. Ceribell has been validated in multiple hospital settings for rapid detection of cerebral dysfunction and has received Breakthrough Device Designation for delirium monitoring, reflecting its clinical potential for use in our study.^25–27^

**Figure 1.**
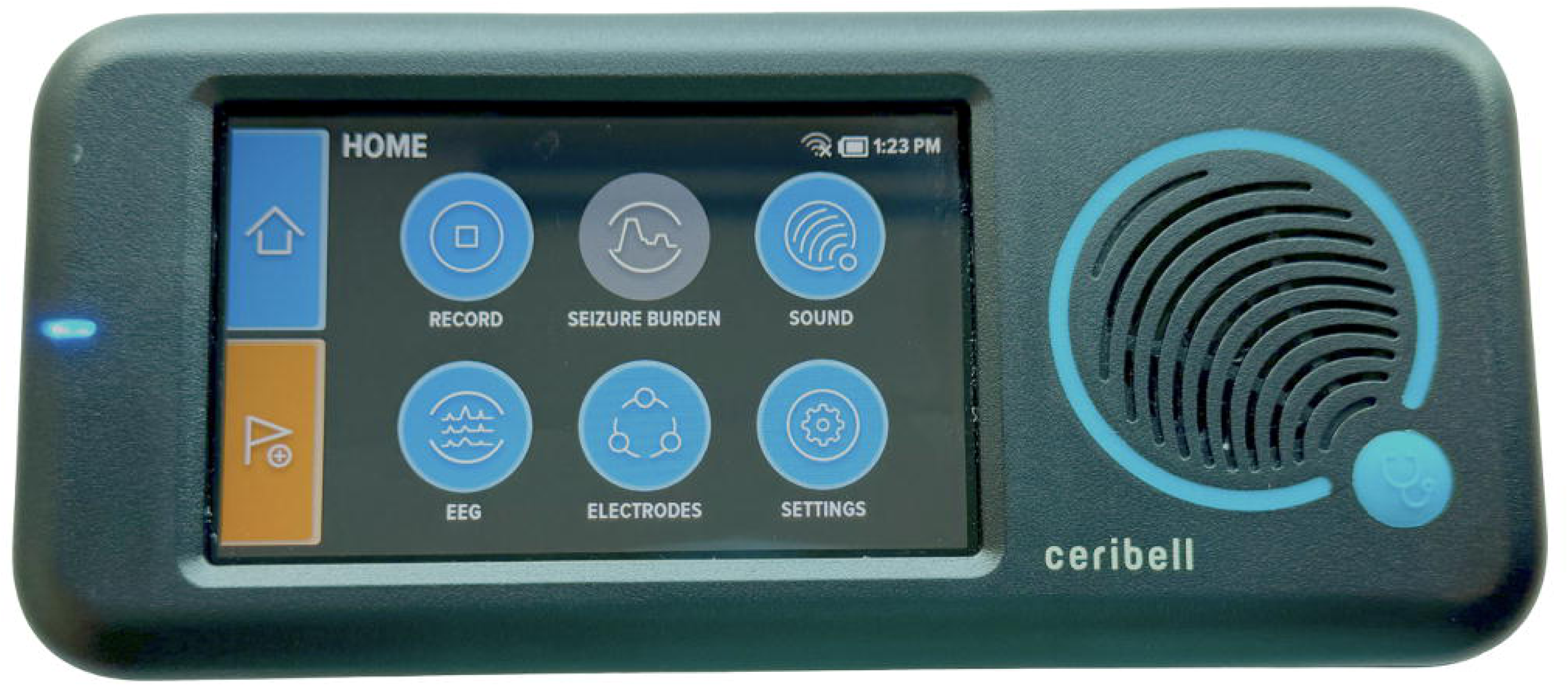
Ceribell EEG system used in the study, including the (A) handheld recorder and (B) disposable ten-electrode headband.

EEG recordings are obtained during five predefined perioperative periods. First, a minimum 15-minute preoperative baseline is recorded to characterize cortical activity at rest, as occipital EEG alpha power during this period has been associated with vulnerability to POD.^28^ Second, continuous EEG acquisition continues as tolerated through anesthetic induction and third, through intraoperative maintenance, during which burst-suppression burden is quantified as a marker of anesthetic depth and potential cerebral stress.^29^ Fourth, the reversal phase corresponding to emergence from anesthesia after extubation is recorded. Finally, recovery in the post-anesthesia care unit (PACU) is then monitored to evaluate the return of alpha activity, which may reflect neurophysiological resilience or early dysfunction.^30^ If visual inspection of PACU EEG data shows poor quality, overnight EEG will be recorded during the first postoperative night to characterize sleep continuity and delta-band activity.^31^

### EEG Acquisition Procedure

Our EEG acquisition procedure follows a standardized protocol between patients. De-identified patient information is first entered into the Ceribell recorder device for organization of data storage. The disposable headband is then sized according to the patient’s estimated head circumference and applied to ensure optimal electrode placement. Conductive gel capsules within the electrodes are activated, and impedance is observed via the EEG recorder touchscreen. Electrodes are manually adjusted until favorable impedance thresholds (≤10 kΩ) are achieved on at least eight of the ten channels and acceptable thresholds (≤30 kΩ) are achieved on the remainder.

After proper EEG setup and completion of the MoCA, preoperative baseline data recording begins immediately. Patients are encouraged to spend at least 15 minutes with their eyes closed, engaging in minimal conversation to obtain a standardized portion of preoperative baseline recordings to compare between patients. Continuous EEG is then maintained as the patient enters the operating room (OR) and anesthesia induction occurs, up through skin closure. Perioperative events such as induction drug administration, incision, closure, and extubation are precisely recorded in the patient’s electronic health record and correlated with the timing recorded on the EEG tracing. Recordings continue for at least 45 minutes after emergence in the PACU **(Figure 2)**. Of note, the device may be temporarily removed or adjusted to accommodate surgical requirements or patient comfort, and impedance is rechecked accordingly after each adjustment. Additional gel can be manually applied between the electrode surface and patient skin via syringe to facilitate acceptable impedance after readjustments. All data are automatically streamed or manually uploaded via secure, encrypted transmission to the Ceribell EEG portal, and European Data Format (.edf) files are provided to the team by Ceribell for subsequent analysis.

**Figure 2.**
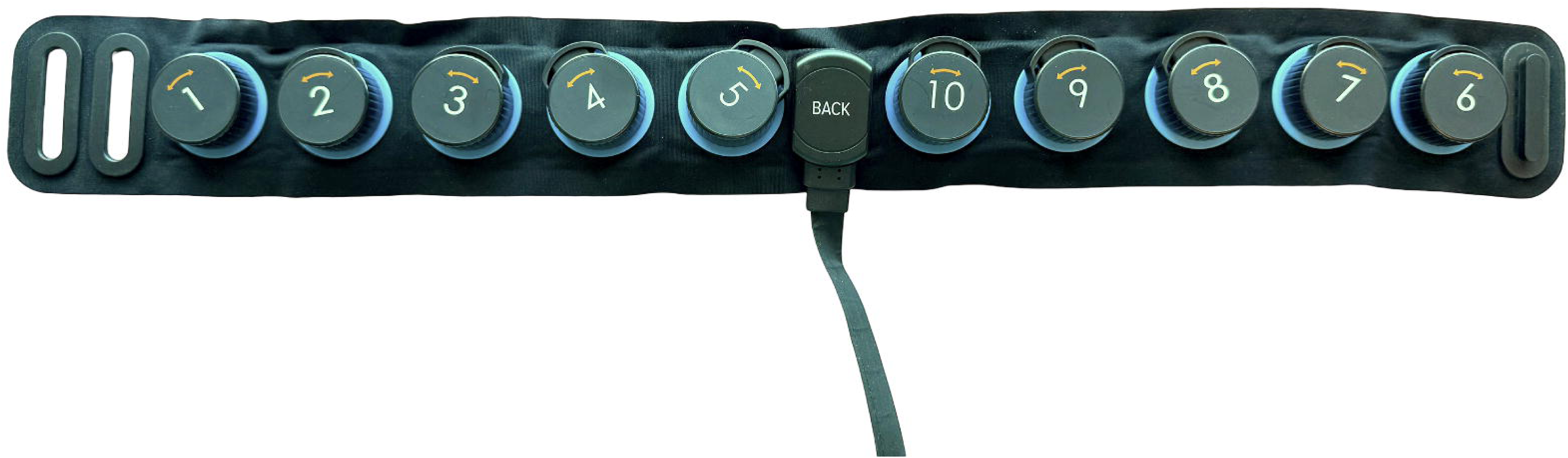
Ceribell EEG system in process of active signal recording.

### Primary Outcome: Delirium Assessment

Delirium is assessed throughout the postoperative period using a structured, tiered approach. Trained nursing staff administer the 4AT delirium screening tool after the patient regains consciousness in the PACU and then every 12 hours during the admission as part of routine care.^32^ Participants who screen positive are subsequently evaluated by a member of the research team using the Confusion Assessment Method-Severity (CAM-S) test, which provides a standardized measure of delirium presence and severity. Delirium outcomes are recorded in the electronic health record for use in model training.

### Secondary Outcomes: Cognitive Assessment and Pain Assessment

Cognitive function is assessed from preoperative baseline through follow-up using the MoCA. The MoCA will be repeated at one day after surgery and at six-week follow up. If in-person testing is not feasible, telephone alternatives (T-MoCA) will be used. Scores are age and education adjusted and summarized to quantify change from baseline. Cognitive outcomes are recorded in the study database for use in secondary analyses and model development.

Although postoperative pain is not hypothesized as a causal determinant of POD, postoperative pain intensity may reflect neural mechanisms of arousal and cortical network dysfunction measurable by neuromonitoring.^33^ By exploring POC EEG correlates of pain along with POD and cognition, we aim to better characterize shared neurophysiological signatures between these perioperative outcomes. Postoperative pain is assessed by bedside nursing staff obtain 0-10 NRS scores immediately before surgery and then every three to six hours while the patient is awake following surgery, per standard nursing protocols, throughout the duration of the patient’s admission. Analgesic administrations are time-stamped and converted to oral morphine milligram equivalents (MME) to quantify exposure, and research staff derive summary measures including pain area-under-the-curve (AUC), time to adequate control (two consecutive NRS ≤4), severe pain burden (proportion of NRS ≥7), and total MME/kg over 48-72 hours. Pain outcomes are abstracted from the electronic health record and study database for secondary analyses and model training.

### EEG Data Analysis

Sample EEG tracings from patients enrolled in the study are presented in **Figure 3**. Per device specifications, signals are digitized at 250 Hz sampling rate with a 0.5-100 Hz hardware bandwidth.^34^ During acquisition, the device automatically measures electrode impedance once per minute using a test signal outside the EEG frequency band to avoid recording artifacts. Any 60-second epoch with impedance values exceeding 30 kΩ will be excluded from analysis. In accordance with prior literature, EEG signals will be band-pass filtered with cutoffs of 0.5 Hz and 50 Hz to filter baseline noise and high-frequency noise.^35^ EEG data will be further screened by an experienced neurologist to ensure artifact rejection. After preprocessing, the clean EEG signals will be segmented into non-overlapping 60-second epochs. This length of time was chosen to balance time resolution with feature estimation and aligns with the device’s 60-second impedance checks.

**Figure 3.**
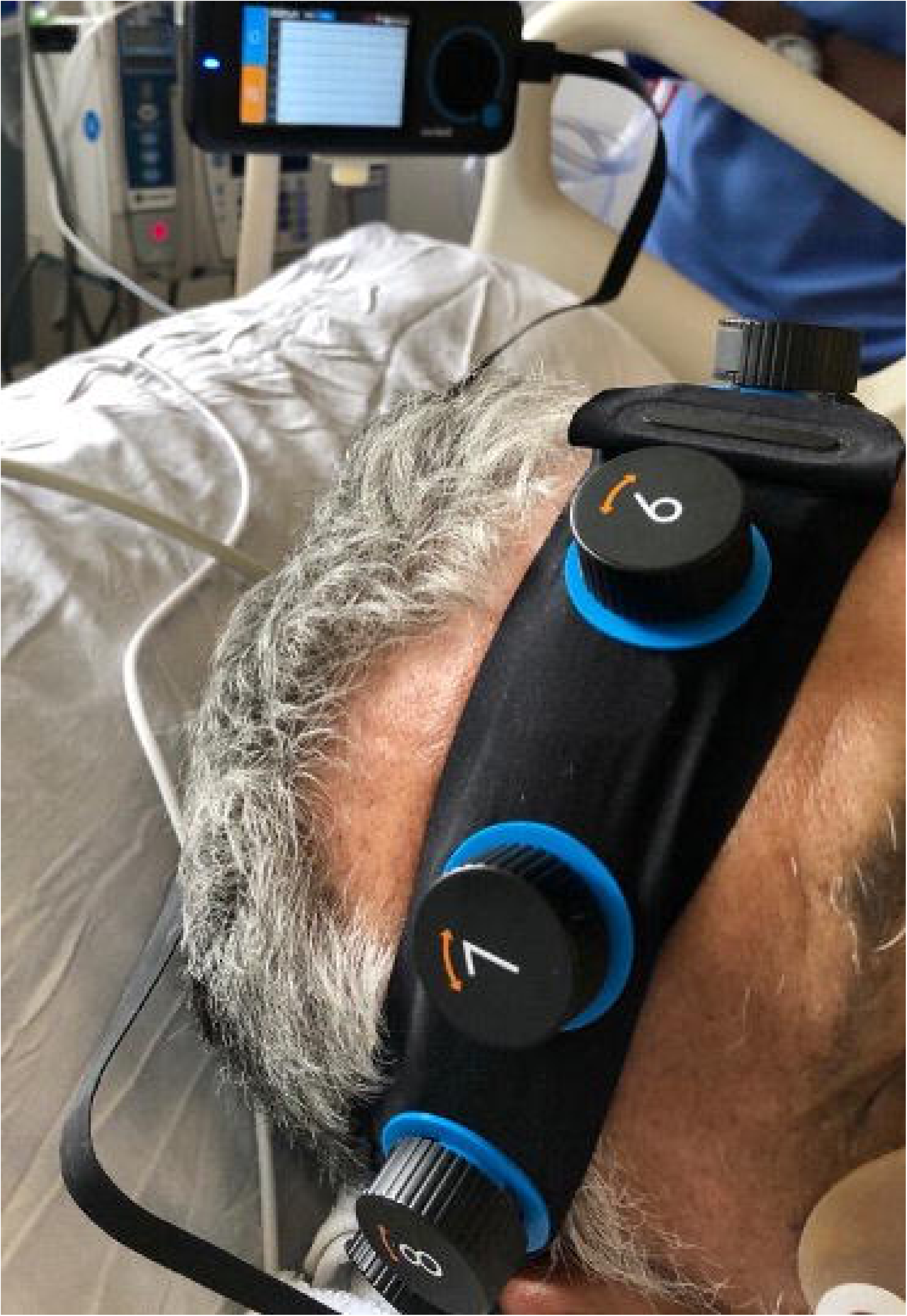
(A) 5-second, (B) 10-second, and (C) 60-second epochs of Ceribell EEG recordings taken at the time of anesthesia induction for patients in the study. The pink markers indicate the moment that the induction agent was administered.

After preprocessing, feature extraction will be conducted in Python (version 3.11) using the MNE-Python library for signal analysis,^36^ along with NumPy and SciPy for numerical computation. For each epoch, the power spectral density is estimated using Welch’s method, and spectral power was integrated within canonical bands (delta 0.5-4 Hz, theta 4-8 Hz, alpha 8-13 Hz, beta 13-30 Hz, and gamma >30 Hz). Absolute and relative power in each band for every included 60-second epoch are calculated independently across each channel. The alpha/delta power ratio is extracted, as alpha/delta power ratios are a prominent feature associated with delirium risk and we hypothesize lower alpha/delta ratios in those who develop POD.^37–39^

Each power spectral density value will be normalized by dividing the frequency bin’s power by the total power across the epoch, yielding a probability distribution which can then be used to calculate spectral entropy using Shannon’s formula.^40^ This approach has been applied to prior delirium studies assessing anesthetic depth and is a key metric for postoperative recordings.^41–44^ Finally, the burst suppression ratio (BSR) is calculated as an elevated BSR may reflect deep anesthetic suppression and has been linked to POD.^45,46^ The BSR is calculated as the proportion of each 60-s epoch during which the EEG amplitude remained below ±5 µV for at least 0.5 s. Suppressed intervals were identified using the Hilbert transform to generate an amplitude envelope, smoothed with a short median filter, and merged if separated by <200 ms. For each epoch, BSR was computed per channel and then summarized as both mean and maximum values across all channels. Recordings that achieve high fidelity may also be analyzed with continuous EEG data. Sun *et al.* describe a deep learning model using a combination of convolutional and recurrent neural networks that allows for continuous monitoring of level of consciousness and delirium in patients admitted to the intensive care unit.^47^

All EEG recordings are annotated with perioperative time stamps and segmented into discrete phases for analysis. Five primary analytic phases have been identified. The preoperative baseline phase consists of at least 15 minutes of resting-state EEG with eyes closed in a quiet environment, providing a baseline measure of cortical rhythms prior to sedation. The induction phase begins at the time of intravenous propofol administration and is defined as the first 5-10 minutes following induction. The intraoperative maintenance phase spans the duration of the surgical procedure, from skin incision to surgical closure, during which steady-state anesthetic exposure was maintained. The reversal phase encompasses the discontinuation of anesthetic agents and initiation of emergence, including the interval leading up to and immediately following extubation. Finally, the recovery phase generally includes a minimum of 45 minutes of EEG recording during postoperative recovery, capturing the early post-anesthetic period as patients regained wakefulness. For certain analyses, this phase may alternatively be defined more broadly (*i.e.,* extending into the first postoperative night) or omitted entirely based on logistical constraints. Segmenting the recordings into these five standardized epochs allows phase-specific feature extraction and facilitates comparisons of EEG dynamics across the perioperative trajectory. These phases have been chosen for their relevance in prior analyses. For instance, alpha power during induction and maintenance anesthesia has shown associations with POD.^48–50^ Moreover, during anesthesia emergence and in the postoperative setting, trajectories with alpha spindle oscillations are proposed to be associated with delirium incidence.^51^

### Blood Biomarker Data Analysis

As previously described, a subset of patients may agree to provide blood samples which will be collected in cfDNA-stabilizing tubes, processed under standardized pre-analytic conditions, and separated into plasma and serum according to institutional standard operating protocols. Plasma will be promptly aliquoted and stored at -80 °C until further analysis is conducted. At that time, plasma cfDNA will be isolated and quantified, with exploratory analyses of tissue-of-origin signatures based on methylation-informed assays. Proteomic profiling will focus on markers of neuroglial inflammation and cognitive impairment such as IL-6 and apolipoprotein E measured on validated targeted platforms.^17,52^ This integrative approach is supported by recent work in spinal cord injury demonstrating that combining cfDNA with plasma protein markers improves diagnostic accuracy and prognostic performance compared with either modality alone.^53^ These assays will be performed blinded to cognitive outcomes, with quality control procedures applied according to standardized protocols. Analysis will be conducted in collaboration with bioinformatics experts at our institution.

### Predictive Model Generation

The primary endpoint for prediction in this study is the incidence of POD. Prediction models will be developed using patient baseline characteristics (age, sex, comorbidity burden), cognitive function (MoCA score), perioperative EEG features, and, where available, blood biomarker data. Three nested models are planned. Model A will include preoperative features only (*i.e.,* baseline characteristics, cognitive function, and resting-state EEG alpha power). Model B will expand to include intraoperative EEG markers such as burst-suppression burden and recovery of alpha activity in the PACU. Model C will serve as an exploratory framework that incorporates the full perioperative feature set, including postoperative sleep metrics and blood biomarker data.

To maximize data efficiency and ensure robust performance estimates, model development and evaluation will be conducted using repeated stratified k-fold cross-validation. Model stability will be further assessed through nested sampling, and bootstrapping procedures will be applied to generate confidence intervals for model performance metrics. Regularized logistic regression will serve as the primary analytic framework to reduce overfitting given the limited number of events, with secondary comparison to alternative machine learning classifiers if performance criteria are not met. Model performance will be evaluated by discrimination (ROC-AUC), calibration (calibration slope, calibration intercept, Brier score), and classification performance metrics (sensitivity, specificity, positive predictive value, negative predictive value). To enhance interpretability, variable importance will be quantified, and feature contributions visualized using SHAP values, enabling an understanding of how clinical, EEG, and biomarker inputs contribute to POD risk stratification.

### Patient Follow-up

At approximately six weeks after surgery, participants who are available will undergo repeat cognitive assessment at their routine follow-up clinic visit using the MoCA to evaluate longer-term cognitive outcomes. Because this assessment is not feasible for all patients, follow-up MoCA data will be treated as a secondary endpoint and analyzed in an exploratory fashion. Where available, six-week MoCA scores will also be incorporated into extended predictive modeling to examine whether perioperative EEG and blood biomarker features are associated with subacute changes in cognition beyond the immediate postoperative period.

### Expected Deliverables

The study is expected to yield several key outputs. First, we aim to generate an EEG-only risk prediction model for POD with a target discriminative performance ROC-AUC of 0.80 or above. Second, we will produce a feature-importance map highlighting physiologically interpretable markers that contribute to delirium risk. Third, a de-identified feature dataset will be released to facilitate independent external validation by other groups. In addition, exploratory analyses integrating blood-based biomarkers with EEG features are anticipated to generate preliminary multi-modal risk models and provide biological insights into perioperative vulnerability. Finally, the full analysis code and preregistered protocol will be archived in publicly accessible repositories to ensure transparency and reproducibility of the work.

## DISCUSSION

### Study Rationale and Anticipated Impact

POD is prevalent, underdiagnosed, and associated with increased mortality and healthcare expenditures.^2,8^ Therefore, identifying POD as early as possible is critical for favorable outcomes, as this opens the door to proactive prevention strategies. The limitations inherent in current screening methods contribute to diagnostic delays and inadequate management. Our study explores the use of POC EEG to generate objective, real-time risk stratification for older adult patients undergoing elective surgery. This methodology aims to identify neurophysiological alterations that precede the manifestation of overt cognitive symptoms.

As noted previously, identifying high-risk patients prior to delirium onset may allow for timely preventive planning. Such planning could include modification of anesthetics, utilization of physical therapy, and optimization of postoperative medication regimens. For instance, one landmark study previously demonstrated that targeted adjustments to treatment plans for patients at risk of delirium reduced its incidence by approximately 5%, significantly decreased immobility, visual and hearing impairments, and lowered expenditures by $6,341 per case prevented.^54^ Preemptive strategies offer a promising path to reduce the incidence and severity of POD, which will improve patient care and decrease the associated healthcare expenditures.

### Strengths of the Study Design

This prospective study utilizes a multi-center recruitment strategy to increase the generalizability of results across diverse clinical environments. The utilization of an FDA-cleared POC EEG device ensures this approach is both clinically reliable and designed for efficient deployment and continuous monitoring in real-world settings.

Additionally, adopting a multimodal analytical framework that integrates traditional cognitive assessment, quantitative electrophysiology, and inflammatory blood biomarkers can help providers design a personalized, comprehensive treatment plan for each patient. This proposed approach aims to be efficiently used by healthcare providers with different training levels without sacrificing the reliability or accuracy of POD diagnosis.

### Potential Clinical Implications

A significant limitation of prior EEG-based delirium predictions lies in their limited generalizability. Traditional EEG systems are cumbersome, requiring extensive electrode placement by a trained clinical staff, which may take 30-60 minutes on average.^55,56^ In contrast, portable EEG devices such as the Ceribell device used in this study can be applied in 1-2 minutes without significant training and are recorded on a handheld device that can be placed on any nearby surface or attached to the patient’s bed or anesthesia equipment. If validated, this approach could serve as a point-of-care tool that integrates seamlessly into the perioperative workflow, enabling prediction of POD and supporting clinical decisions to mitigate risk. In the context of precision medicine, this approach could serve as a novel perioperative risk-stratification tool, enhancing the ability to anticipate POD and tailor management strategies. When combined with a blood test, real-time EEG signatures and global semi-static blood biomarkers can create a multimodal risk profile incorporating immediate changes in cortical physiology in a background index of neuronal health, inflammation, and systemic stress. Ultimately this research could lay the groundwork for interventional trials where predicted high-risk patients may be more readily identified so that management strategies may be employed.

### Limitations

Several limitations may constrain the generalizability and applicability of integrating portable EEG into perioperative workflows. First, the study’s modest sample size could pose challenges to model accuracy, especially given that POD incidence typically ranges from approximately 17-24% among surgical populations depending on patient demographics and delirium assessment methods.^57^ To this point, our use of the 4AT, which is a subjective delirium assessment tool, may lead to false positive or negative cases that would be used to incorrectly train and evaluate or model. Next, incomplete or missing data such as device disconnection, prolonged high impedance recordings, omitted delirium assessments, lack of blood biomarkers, or patient attrition may introduce bias or reduce statistical power. Although POC EEG’s appeal lies in its mobility and ease of deployment, implementation within two hospitals at a single-institution academic center may reflect an environment uniquely suited to such technology, potentially limiting external applicability.

The Ceribell device itself also presents intrinsic constraints. Originally validated for seizure detection, it utilizes a limited-channel montage, favoring broad spectral features over regional specificity.^58^ These features allow for detection of global patterns but reduce its sensitivity to focal cortical changes associated with delirium. Signal quality concerns, including device misplacement and non-ideal electrode contact, could further affect data reliability. Evaluations of similar POC EEG systems have noted limitations in spatial coverage and signal fidelity when compared to research-grade systems.^59^ Finally, our analysis of blood biomarkers remains exploratory; while these biomarkers may offer valuable insights, they are not diagnostic and do not substitute for clinical delirium assessments.

### Future Directions

Future work should prioritize the expansion of our protocol to larger, multi-center cohorts for further external validation and to ensure generalizability across diverse surgical populations and institutional practices. Furthermore, EEG signatures and biomarker profiles may be utilized to not only predict delirium but long-term cognitive outcomes. Finally, embedding predictive EEG analytics into the perioperative workflow could allow clinicians to adapt anesthetic dosing, implement non-pharmacologic interventions, or prioritize closer postoperative monitoring when high-risk patterns emerge. Ultimately, such interventional trials will determine whether predictive monitoring can be translated into improved outcomes through actionable prevention strategies.

## CONCLUSION

This prospective cohort study aims to evaluate whether a POC EEG device, with or without adjunctive blood-based biomarker data, can provide an accurate and clinically feasible method to predict POD in older patients undergoing elective surgery. By integrating objective neurophysiological signatures with conventional risk factors, this work seeks to move beyond subjective screening toward real-time, administrator-independent prediction. The anticipated outputs, including a validated EEG-based risk model, multimodal exploratory analyses, and openly shared datasets, aim to advance both scientific understanding and practical application of POD risk stratification. If successful, this approach will lay the foundation for interventional strategies that identify high-risk patients preemptively, enabling timely POD preventive measures and ultimately improving perioperative outcomes while reducing healthcare costs.

## TRIAL STATUS

**Protocol Version:** v1 (October 2025)

**Recruitment:** Recruitment began on August 1, 2025 and is planned to be completed by August 1, 2026

## DECLARATIONS

### Ethics Approval and Consent to Participate

This study was approved by the Johns Hopkins University Institutional Review Board (IRB00463945). Written, informed consent to participate is obtained from all participants.

## Author Contributions

NT and TDA are chief investigators who conceived the study, led protocol development, and supervise the work. VNV, PK, and NA are responsible for investigation. VNV, PK, IY, and SVS are responsible for data analysis. ADD provides logistical support. The remaining authors provide methodological support. All authors approved the manuscript.

**Previous Presentations:** None.

**Conflicts of Interest:** None.

## Data Availability

All data produced in the present study will be made available upon reasonable request to the authors

## LIST OF ABBREVIATIONS

AUC: area under the curve
BSR: burst suppression ratio
CAM: Confusion Assessment Method
CAM-S: CAM-Severity
cfDNA: cell-free DNA
EEG: electroencephalography
EPP: events-per-parameter
IRB: Institutional Review Board
MoCA: Montreal Cognitive Assessment
MME: morphine milligram equivalent
NRS: Numeric Rating Scale
NSR: non-significant risk
OR: operating room
PACU: post-anesthesia care unit
POC: point-of-care
POD: postoperative delirium
ROC-AUC: area under the receiver operating characteristic curve
T-MoCA: telephone MoCA
TRIPOD: Transparent Reporting of a multivariable prediction model for Individual Prognosis or Diagnosis

---

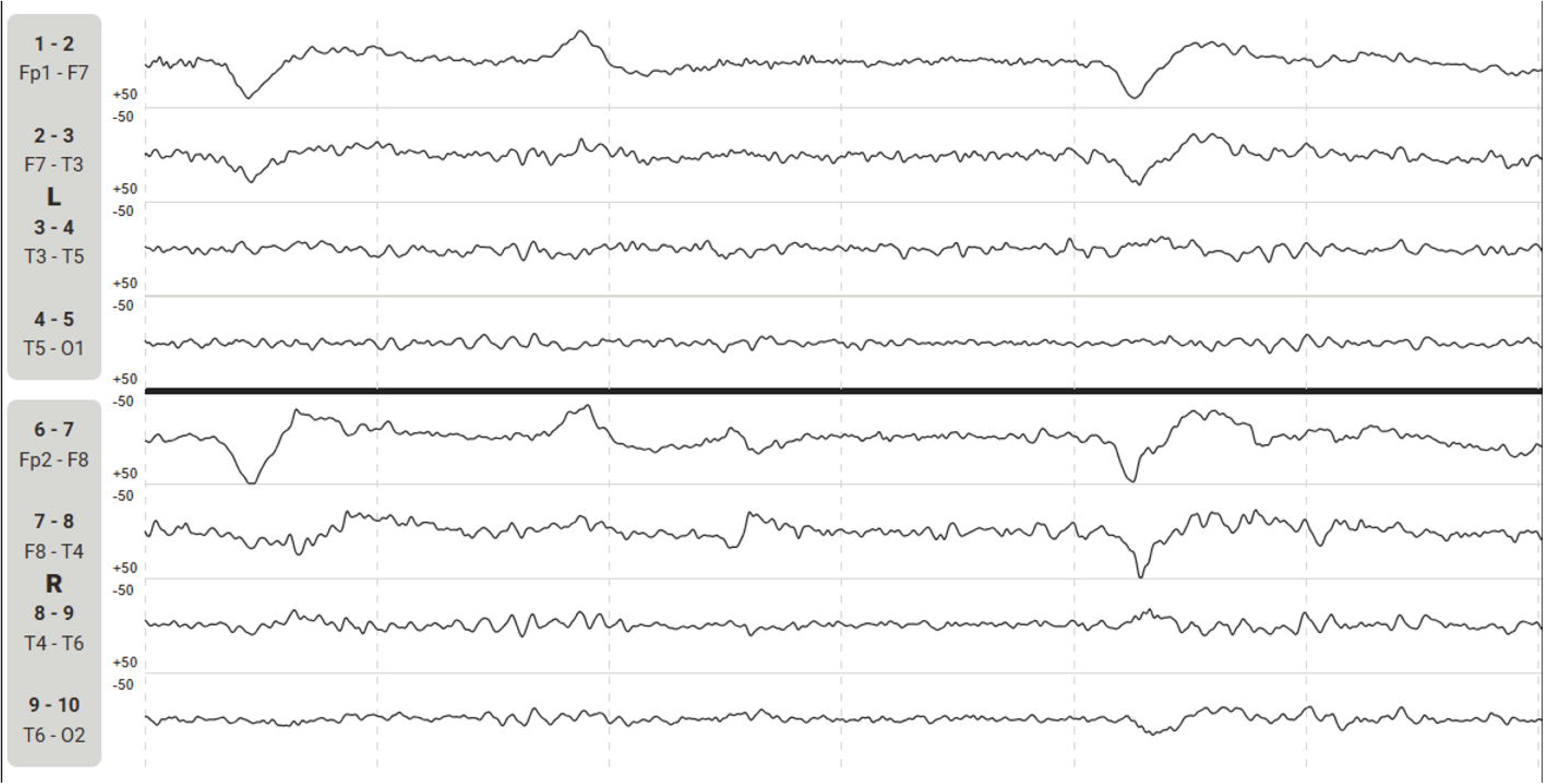

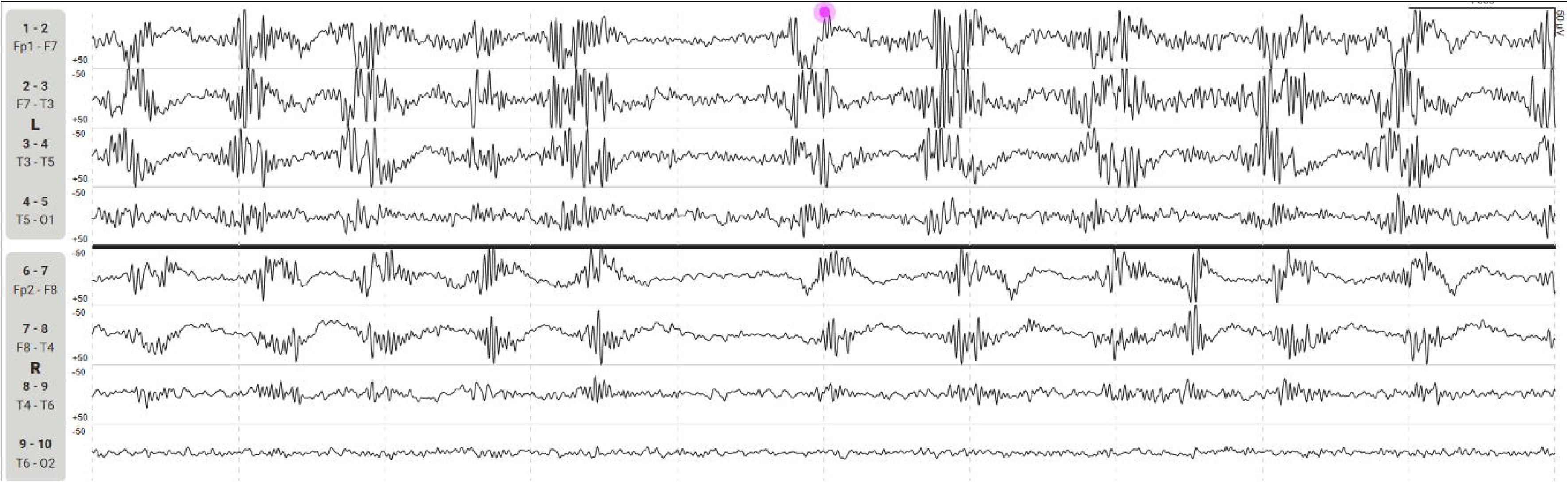

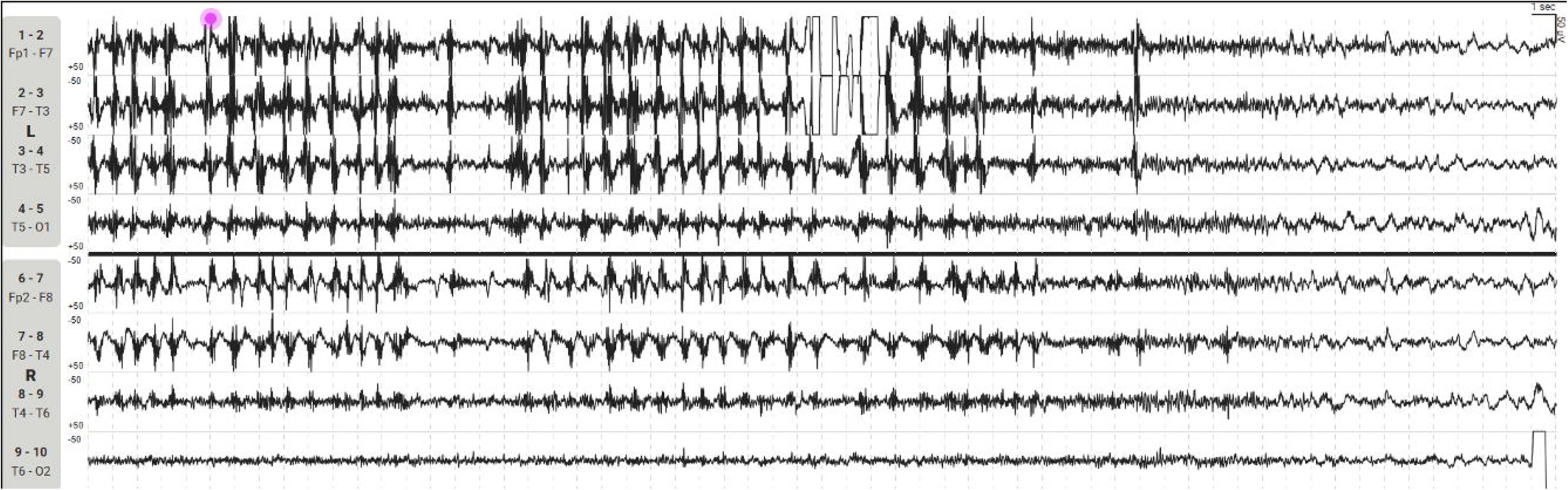

